# Digital gait biomarkers, but not clinical ataxia scores, allow to capture 1-year longitudinal change in Spinocerebellar ataxia type 3 (SCA3)

**DOI:** 10.1101/2022.03.10.22272119

**Authors:** Winfried Ilg, Björn Müller, Jennifer Faber, Judith van Gaalen, Holger Hengel, Ina R. Vogt, Guido Hennes, Bart van de Warrenburg, Thomas Klockgether, Luger Schöls, Matthis Synofzik, the ESMI Consortium

## Abstract

Measures of step variability and body sway during gait have shown to correlate with clinical ataxia severity in several cross-sectional studies. However, to serve as a valid progression biomarker, these gait measures have to prove their sensitivity to robustly capture longitudinal change, ideally within short time-frames (e.g. one year). We present the first multi-center longitudinal gait analysis study in spinocerebellar ataxias (SCAs). We performed a combined cross-sectional (n=28) and longitudinal (1-year interval, n=17) analysis in SCA3 subjects (including 7 pre-ataxic mutation carriers). Longitudinal analysis revealed significant change in gait measures between baseline and 1-year follow-up, with high effect sizes (stride length variability: p=0.01, effect size r_prb_=0.66; lateral sway: p=0.007, r_prb_=0.73). Sample size estimation for lateral sway reveals a required cohort size of n=43 for detecting a 50% reduction of natural progression, compared to n=240 for the clinical ataxia score SARA. These measures thus present promising motor biomarkers for upcoming interventional studies.

## Introduction

With disease-modifying drugs on the horizon for degenerative ataxias^1, 2^, sensitive motor biomarkers are highly warranted. Gait variability measures including step variability and body sway have shown their sensitivity to ataxia severity in multiple cross-sectional studies, correlating with clinical ataxia scores^3-15^. However, correlations with ataxia scores in cross-sectional studies are strongly influenced by the range of disease severity (range of observations^16^): For cohorts that encompass a wide range of disease stages, many gait measures - including unspecific ones like gait speed-show significant correlation to disease severity, often predominantly driven by subjects at the ends of the spectrum of disease severity^16^. In interventional trials, the goal of assessing motor biomarkers is qualitatively different: namely the quantification of individual change in short time-frames (e.g. one year).

To serve as valid progression biomarkers, these gait measures thus have to prove their sensitivity to individual longitudinal change in a sufficiently short time-span realistic for intervention trials. Here, we present a first longitudinal study in a multi-center SCA cohort. We demonstrate that digital-motor biomarkers allow to capture longitudinal change with high effect sizes within just 1-year follow-up, with this sensitivity to change outperforming clinical ataxia scores.

## Methods

### Patients

The study cohort was part of the European Spinocerebellar ataxia type 3/Machado-Joseph disease initiative (ESMI), a large multi-centre prospective observational study. 28 mutation carriers of Spinocerebellar Ataxia type 3 (SCA3) were recruited from the Ataxia Clinics of the University Hospitals Tübingen and Nijmegen as well as the German Center for Neurodegenerative Diseases (DZNE) in Bonn. They comprised of 21 subjects at the ataxic stage as defined by a Scale for the Assessment and Rating of Ataxia (SARA)^17^ score of ≥3 (subgroup SCA3_ATX_; mean SARA 8.37 points), and 7 subjects at the pre-ataxic stage (SARA score <3) (subgroup SCA3_PRE_; mean SARA 1.08 points). Neurological signs other than ataxia were assessed by the Inventory of Non-Ataxia Signs (INAS)^18^. Healthy controls (N=13, SARA 0.9±0.7, age 42.3±15) comprised mutation-negative first-degree relatives of SCA3 carriers and unrelated healthy individuals, all without symptoms or signs of neurodegenerative disease.

The study was approved by the local institutional review boards of all participating centres. Written informed consent was obtained from all study participants before enrolment.

### Gait assessment

Gait was recorded at each study site by a multi-Kinect recording system with six cameras (for detailed description, see Supplement 1 and^19^). In a previous validation study^19^, this system has shown to deliver good-to-excellent accuracy in several gait measures, including step length und step duration.

We assessed gait movements in two conditions: preferred speed and slow speed. The slow speed condition was included based on previous studies demonstrating increased step variability and body sway for slow gait^20^. In both conditions, subjects walked in their usual everyday life shoes a 10m distance for 5 trials. Out of all potential gait parameters, we here chose a hypothesis-based approach focussing on measures that were considered as promising candidates in degenerative ataxia based on previous studies^3, 10, 13, 15, 21^, namely measures on step variability and lateral body sway. Variability measures were calculated using the coefficient of variation CV=σ/µ, normalizing the standard deviation with the mean value^22^. As measures of step variability, stride length CV (StrideL_CV_) and stride time CV (StrideT_CV_) were determined. Lateral body sway was defined as the medial–lateral component of the path of the sternum marker (see Supplement 1), normalized by the anterior–posterior component^9^.

### Statistics

Between-group differences were determined by the non-parametric Kruskal-Wallis-test. When the Kruskal-Wallis-test yielded a significant effect (p<0.05), post-hoc analysis was performed using a Mann-Whitney U-test. Effects sizes were determined by Cliff’s delta^23^. Repeated measurements analyses were performed for longitudinal analyses using the non-parametric Friedman-test to determine within-group differences between assessments. When the Friedman-test yielded a significant effect (p<0.05), post-hoc analysis was performed using a Wilcoxon-signed-rank-test for pairwise comparisons. Effect sizes for the repeated measurements analyses were determined by matched-pairs rank biserial correlation^24^. Estimated time to ataxia onset was calculated based on individual’s CAG repeats, as described in ^25^. We report three significance levels: (i) uncorrected *:p<0.05: (ii) Bonferroni-corrected for multiple comparisons **:p<0.05/n with n=6: number of analysed gait features including both walking conditions; (iii) ***:p<0.001. Spearman’s ρ was used to examine the correlation between gait measures and SARA scores. Statistical analysis was performed using MATLAB (Version R2020B). Based on the longitudinal changes, a sample size estimation was performed using G*power 3.1^26^ to determine the required cohort size for different levels of reduction of natural progression by a hypothetical intervention.

## Results

### Correlation of gait measures to cross-sectional ataxia severity

Cross-sectional analysis revealed significant group differences between SCA3 vs HC in all examined gait measures in both walking conditions (e.g. lateral sway in preferred speed: p=0.00011; slow speed p=0.003, Table 1, Figure 1A). Step variability measures and lateral sway showed significant relationship to cross-sectional ataxia severity in both walking conditions, with highest effect sizes for stride length CV (δ=0.64) (Table1). For slow walking, step variability measures (p=0.016) and lateral sway (p=0.043) also correlated with SARA in the subgroup of ataxic SCA3 mutation carriers (SCA3_ATX_).

**Table 1.** Results of cross-sectional and longitudinal analyses. Cross-sectional analyses: Between-group differences of healthy controls (HC) and SCA3 subjects for clinical measures and gait measures in the walking conditions with preferred and slow speed. Stars indicate significant differences between groups (*≡ p<0.05, **≡ p<0.0083 Bonferroni-corrected, ***≡ p<0.001). δ denote the effect sizes determined by Cliff’s delta. Correlations between gait measures and clinical ataxia severity (SARA total score, SARA_p&g_ posture&gait subscore) are given for the SCA3 group. The 3 items of the SARA assessing gait and posture (gait, stance, sitting) were grouped by the subscore SARA posture ɖ gait (SARA_p&g_)^21, 35^. Effect sizes of correlations are given using Spearman’s ρ. **Longitudinal analyses of 1-year follow-up assessments:** Paired statistics for within-subject comparisons of clinical scores and gait measures for the two walking conditions (p-values, Wilcoxon signed-rank test; effect sizes r_prb_ determined by matched pairs rank biserial correlation ^24^). m: mean; sd: standard deviation. Shown are analyses for the group of SCA3 subjects at baseline (**SCA3**^**BL**^) and 1-year follow-up (**SCA3**^**FU**^).

|  | Cross-sectional analyses |  |  |  |  |  | Longitudinal analysis |  |  |  |
| --- | --- | --- | --- | --- | --- | --- | --- | --- | --- | --- |
|  | group difference<br>SCA3 vs. HC |  | correlations SCA3 |  |  |  | SCA3<sup>BL</sup> | SCA3<sup>FU</sup> | Within group<br>difference<br>SCA3<sup>BL</sup> vs. SCA3<sup>FU</sup> |  |
|  |  |  | SARA |  | SARA<sub>p&g</sub> |  |  |  |  |  |
|  | p | δ | ρ | p | ρ | p | m ± sd | m ± sd | p | r<sub>prb</sub> |
| Clinical scores |  |  |  |  |  |  |  |  |  |  |
| SARA | 0.0001*** | 0.77 | - | - | - | - | 5.32±4.73 | 5.91 ± 4.91 | 0.2 | 0.27 |
| SARA<sub>p&g</sub> | 0.0002*** | 0.69 | - | - | - | - | 2.12±2.2 | 2.47 ± 2.43 | 0.125 | 0.40 |
| INAS | - | - | - | - | - | - | 2.31±1.53 | 2.81 ±2.19 | 0.138 | 0.44 |
| Walk – preferred speed |  |  |  |  |  |  |  |  |  |  |
| Stride duration CV | 0.10 | 0.32 | 0.18 | 0.36 | 0.33 | 0.084 | 0.064±0.06 | 0.062±0.02 | 0.26 | 0.32 |
| Stride length CV | 0.04* | 0.4 | 0.54 | 0.003** | 0.64 | 0.0003*** | 0.097± 0.1 | 0.129 ± 0.11 | 0.06 | 0.5 |
| Lateral sway | 0.0011 ** | 0.67 | 0.26 | 0.19 | 0.27 | 0.17 | 0.12 ± 0.06 | 0.144 ± 0.07 | 0.01* | 0.68 |
| Walk – slow speed |  |  |  |  |  |  |  |  |  |  |
| Stride duration CV | 0.02* | 0.48 | 0.5 | 0.007** | 0.45 | 0.016* | 0.08±0.03 | 0.1±0.04 | 0.63 | 0.17 |
| Stride length CV | 0.015* | 0.52 | 0.46 | 0.014 * | 0.53 | 0.004** | 0.087±0.03 | 0.103±0.04 | 0.01* | 0.66 |
| Lateral sway | 0.003 ** | 0.63 | 0.54 | 0.002** | 0.52 | 0.021* | 0.128± 0.03 | 0.151 ± 0.04 | 0.007** | 0.73 |

**Figure 1.**
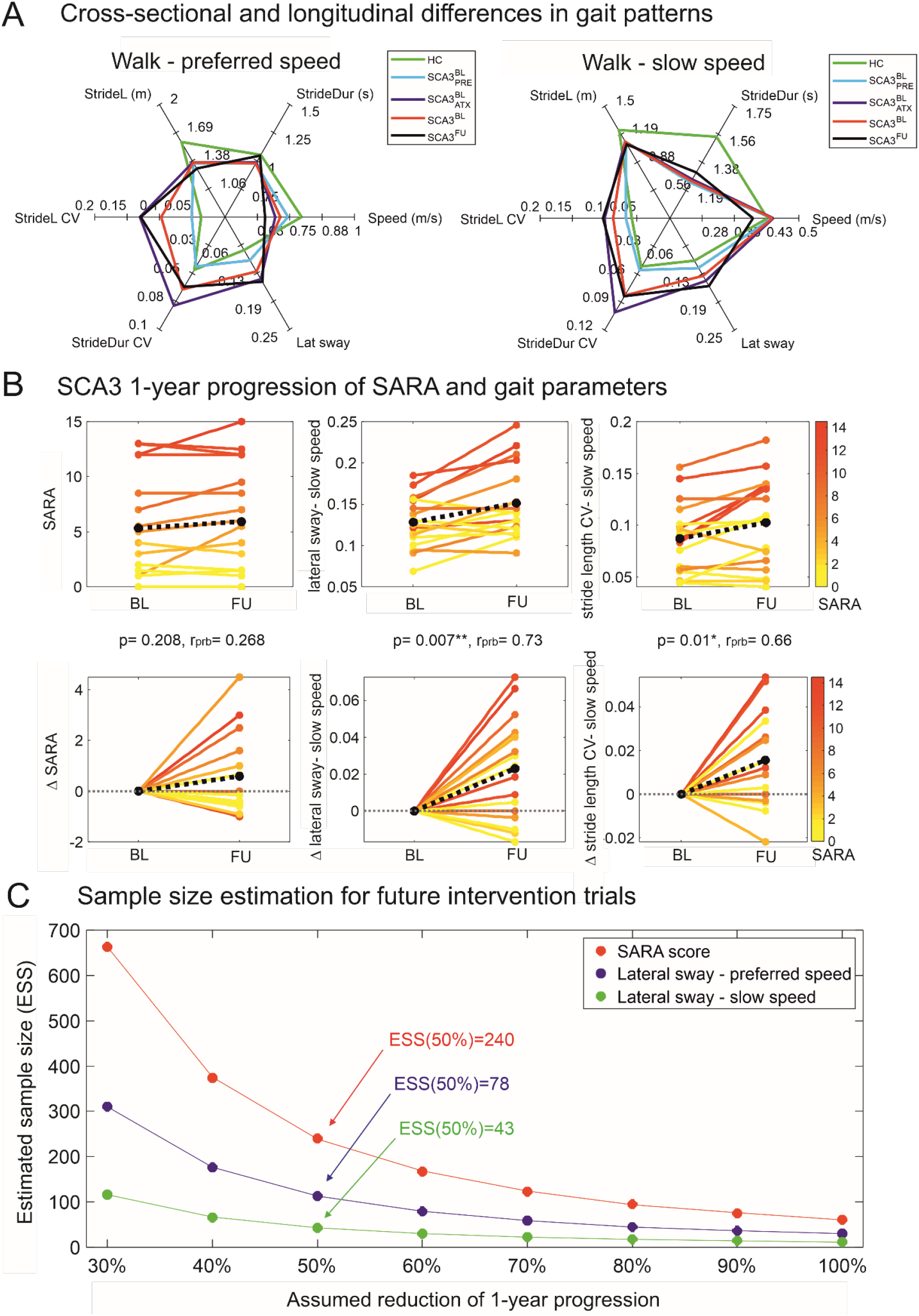
(A) Radar plots illustrating cross-sectional and longitudinal differences on six gait parameters for the two gait conditions with preferred speed and slow speed: Gait speed, Stride Duration (StrideDur), Stride Length (StrideL), Stride Length variability (StrideL-CV), Stride Duration variability (StrideDur-CV), Lateral Sway (Lat sway). Cross-sectional differences can be seen by comparison of healthy controls (HC, green), the subgroup of SCA3 pre-ataxic mutation carriers (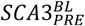, light blue), ataxic mutation carriers (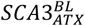, purple) as well as the total group of SCA3 subjects (SCA3^BL^, red). Given are average values for each group. Longitudinal 1-year progression can be seen comparing SCA3 subjects at baseline (SCA3^BL^, red) and 1-year follow-up (SCA3^FU^, black). (B) Longitudinal analyses of 1-year follow-up assessments: Within-subject changes between baseline and 1-year follow-up for the group of SCA3 subjects. Upper panel: Within-subject changes of the SARA score and the gait measures lateral sway and Stride length CV in the slow walking condition at baseline (BL) and 1-year follow-up (FU). Lower panel: Within-subject changes between baseline and 1-year follow-up represented as delta (Δ). In all panels, SARA scores of individual cerebellar subjects are colour coded. Black dotted line = mean change across all subjects. Stars indicate significant differences between timepoints (*≡ p<0.05, **≡ p<0.0083 Bonferroni-corrected, ***≡ p<0.001). Effect sizes r_prb_ were determined by matched-pairs rank biserial correlation. (C) Sample size estimations were performed for future intervention trials showing different levels of reduction in progression levels for the different outcome measures: SARA, lateral sway in the walking conditions with preferred and slow speed. The estimated number of subjects per study arm is plotted over the assumed therapeutic effect for lowering the 1-year progression in SCA3 subjects.

### Sensitivity of gait measures to longitudinal change after one year

We next analysed whether gait measures allow to detect longitudinal changes at 1-year follow-up assessment (duration: 377±33 days). 1-year follow-up data were available from 17 subjects SCA3^FU^ (10 ATX, 7 PRE). Reasons for drop-out from longitudinal recording were unavailability for follow-up assessment (n=6), technical problems in follow-up assessment (n=4), and disability in walking without walking aids at follow-up due to disease progression (n=1).

While the SARA score (mean baseline: 5.32, follow-up: 5.9, p=0.2, effect size r_prb_=0.27) as well as the INAS score (mean baseline: 2.12, follow-up: 2.47, p=0.125, r_prb_=0.4) failed to detect longitudinal change (Table 1, Figure 1B), paired statistics revealed differences between baseline and follow-up in gait measures for both walking conditions (*preferred speed*: lateral sway, p=0.01*, r_prb_=0.68; *slow speed*: lateral sway, p=0.007*, r_prb_=0.73, stride length cv: p=0.01*, r_prb_=0.66, Table 1, Figure 1A+B).

Given the highest effect size from these gait measures, lateral sway was selected for sample size calculation. For detecting a 50% reduction of natural progression by a hypothetical intervention (95% power and one-sided 5% type I error), n=43 subjects would be required if taking the lateral sway during slow walking as primary outcome measure - compared to n= 240 subjects if taking the SARA as primary outcome measure (Figure 1C).

Subgroup analyses revealed even higher effect sizes in sensitivity to longitudinal change for the ataxic SCA3 subgroup 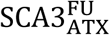 (preferred speed: lateral sway, p=0.03*, r_prb_=0.74; slow speed: lateral sway, p=0.01*, r_prb_=0.87), indicating that the high sensitivity to change is predominantly driven by the ataxic subjects SCA3_ATX_ (Supplement-S2).

## Discussion

Gait disturbance often presents as the first sign of degenerative cerebellar ataxia^27, 28^ and is one of the most disabling features throughout the disease course; thus suggesting a high potential as both progression and response marker in upcoming treatment trials^1, 2, 29^.

This study aimed to test the sensitivity of gait measures to capture longitudinal change within 1 year in a SCA population (SCA3) and a multi-center setting. Analyses revealed that gait measures (i) correlate with cross-sectional clinical ataxia severity, thus indicating valid capture of clinical ataxia dysfunction; and in particular (ii) capture longitudinal change between baseline and 1-year follow-up with high effect sizes, hereby substantially outperforming clinical ataxia scales.

### Gait measures are sensitive to cross-sectional ataxia severity

Measures describing the variability of gait pattern have proven cross-sectional sensitivity to ataxia severity in a wide range of recording methods^30, 31^ including marker-based capturing systems as gold standard^9, 10, 21^, gait mattresses^13, 20^, wearable inertial sensors^4, 5, 7, 8^, and camera-based systems^6^ like the multi-kinect system^19^.

Our results validate these findings in a multi-center setting and in an early stage SCA3 cohort (low SARA score with mean 8.1 points), thus demonstrating their applicability to early disease stages of SCA, as targeted by upcoming interventional studies.

### Gait measures capture longitudinal change with high effect size within one year

However, it is key for upcoming interventional trials that sensitivity to change of these gait markers is proven by quantification of individual changes in short, trial-like time-frames. For upcoming disease-modifying drugs in SCA, the main outcome will be slowing of disease progression in a limited study period, ideally capturable already within e.g. 1 year.

Our SCA3 cohort presents a paradigmatic example for upcoming interventions as SCA3 is the globally most frequent SCA genotype with relatively fast progression^32, 33^ and first interventional trials are expected still in 2022. In our cohort we observed a smaller annual change in the SARA total score than reported for SCA3 in earlier studies^32, 33^ (0.6 vs. 1.56 in^32^), which is most probably due to the earlier disease stage at baseline in our study (mean SARA 5.57 vs. 14.1 in^32^), thus also representing better the disease strata included in upcoming SCA3 intervention trials.

Whereas neither annual change in the ataxia score (SARA) nor in non-ataxia items (INAS) reached significance (Table 1), gait measures captured progression in stride length variability and lateral body sway in both walking conditions, especially in slow walking. The large effect sizes in these gait measures lead to substantially reduced sample size estimations in comparison to the SARA score for the detection of decreased disease progression within one year (Figure 1C). This reduction in sample size is actually decisive whether a trial is feasible at all: while trials with e.g. 240 SCA3 subjects (as required for SARA as outcome) are almost not possible, 43 SCA3 subjects (as required for the gait biomarkers) are well feasible.

### Limitations of the study

Our findings are limited by the relatively small cohort size. In particular, our study cohort was not sufficiently powered for detecting longitudinal change within the pre-ataxic group only. Thus, larger future studies are needed, including in particular a higher number of pre-ataxic subjects, to further validate the promises of gait measures. In future multi-center studies, the gait measures identified here might also be assessed not by a stationary analysis set-up, but rather by wearable inertial sensors^4, 5, 8^, which would -as important for ecological relevance-also allow assessments of these measures in patients’ real life^4, 34^.

## Conclusion

Our study demonstrates that digital gait measures allow to capture natural progression in SCA3 within just one year, with effect sizes outperforming clinical rating scales as the main established outcome measures in the field. They thus present promising motor biomarkers for upcoming SCA intervention studies.

## Data Availability

. Data will be made available upon reasonable request and as patient consent
allows. The authors confirm that the data supporting the findings of this study are available within
the article and its Supplementary material.

## Author Roles

1. Research project: A. Conception, B. Organization, C. Execution;
2. Statistical Analysis: A. Design, B. Execution, C. Review and Critique;
3. 3.Manuscript Preparation: A. Writing of the first draft, B. Review and Critique;

W.I.: 1A, 1B, 2A, 3A

B.M.: 1C, 2C, 3B

J.F.: 1B, 1C, 3B

J.vG.: 1B, 2C, 3B

H.H.: 1C, 2C, 3B

I.V.: 1C, 2C, 3B

G.H.: 1C, 2C, 3B

B.vW.: 1A, 2C, 3B

T.K.: 1A, 2C, 3B

M.S.: 1A, 2C, 3A

L.S.: 1A, 2C, 3B

## Supplementary Information

Supplement 1: The multi-Kinect system

Gait movements were assessed at each site by a multi-Kinect recording system with six cameras (Figure S1-1, see ^19^ for details of hardware and software architecture). After recording, we used the Kinect SDK v2 for skeleton fitting and customized MATLAB routines for merging the six skeletons. Foot events and gait cycles were semi-automatically determined using foot markers and angle trajectories from the skeleton. Trials were smoothed with a Savitzky-Golay polynomial filter and resampled equidistantly with 101 data points per gait cycle by linear time interpolation. In a previous validation study^19^, this system has shown to deliver good-to-excellent accuracy in several gait measures including stride length and stride duration. Details on hardware and software architecture as well as validation with a Vicon system as gold standard can be found in^19^.

**Figure S1-1.**
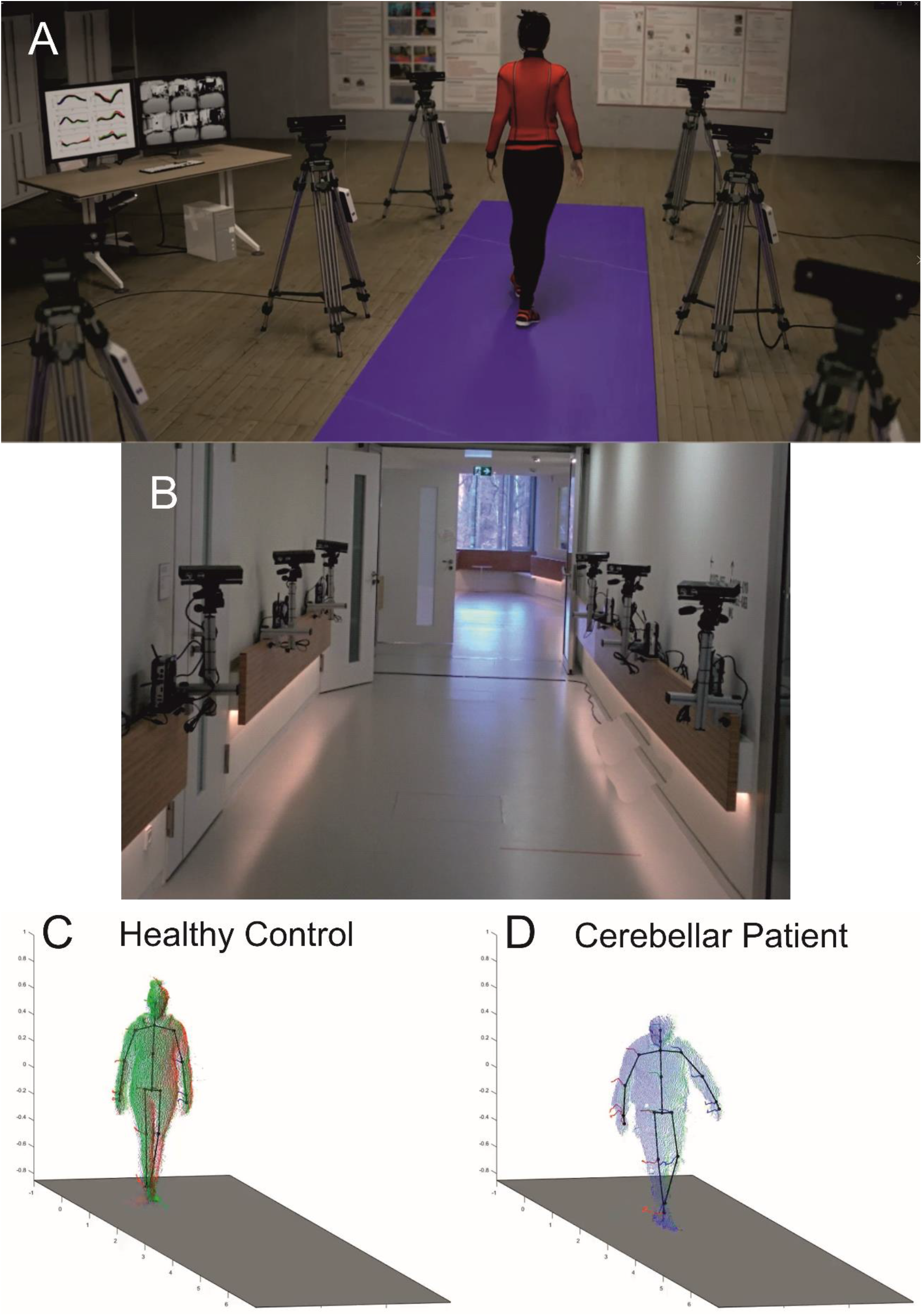
Illustrations of the configuration and resulting output of the multi-Kinect capturing system^19^. (A) An animated view of the overall capturing system including 6 synchronised Kinect v2 cameras. (B) Setup used in Bonn, at the in German Center for Neurodegenerative Diseases (DZNE). (C+D) Snapshot of the resulting output consisting of skeleton trajectories. Colour coding of underlying point clouds is according to the different Kinect systems which delivered data at this time point.

## Supplement S2 – Follow-up results for the ataxic SCA3 subgroup SCA3ATX

**Table S3-1.**
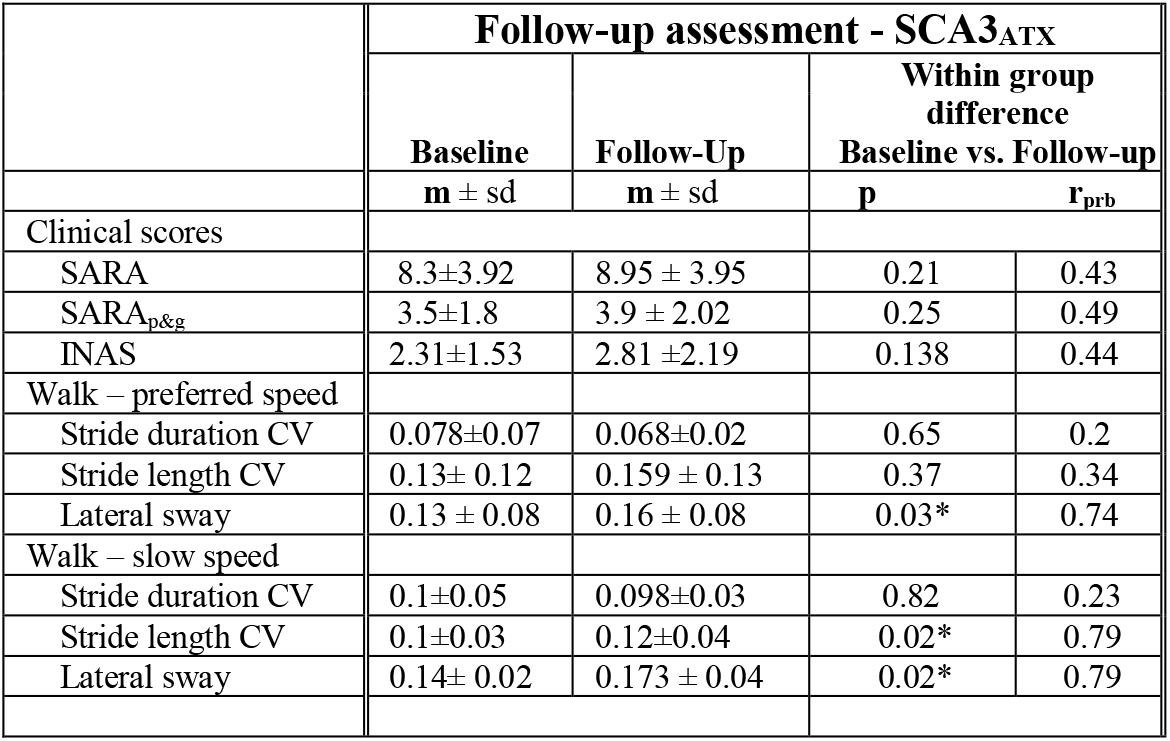
Results of longitudinal analyses of 1-year follow-up assessments for the SCA_ATX_ group. Paired statistics for the within-subject comparison of clinical scores (SARA total score, SARA_p&g_ posture&gait subscore, INAS score) and gait measures for the two walking conditions (p-values, Wilcoxon signed-rank test; effect sizes r_prb_ determined by matched pairs rank biserial correlation ^24^). Shown are analyses for the subgroup of ataxic SCA3 subjects (SCA3_ATX_). Stars indicate significant differences between groups (*≡ p<0.05, **≡ p<0.016 Bonferroni-corrected, ***≡ p<0.001).

